# *“No official help is available”* - experience of parents and children with congenital heart disease during COVID-19

**DOI:** 10.1101/2020.07.03.20146076

**Authors:** LV Marino, R Wagland, DJ Culliford, T Bharucha, SC Sodergren, AS Darlington

## Abstract

**Purpose of the study:** The purpose was to explore the experience, information, support needs and decision-making of parents with congenital heart disease (CHD) during the COVID-19 crisis.

**Study design:** On-line survey design

**Setting:** An on-line survey with open/closed questions to explore the experiences of parents of children, as well as children and young people (CYP) with CHD during the COVID-19 crisis

**Patients:** Parents of children with CHD and CYP

**Results:** 184 parents and 36 CYP completed the survey. Parents worry about the virus (86.4%) vs. CYP (69.4%), whilst (89%) parents are vigilant for symptoms of the virus vs. CYP(69.4%). A thematic analysis of the qualitative comments covered 34 subthemes, forming eight-overarching themes: *Virus*:1)risk of infection, 2)information, guidance and advice, 3)change in health care provision, and 4)fears and anxieties; *Lockdown and isolation*: 5)psychological and social impact, 6)keeping safe under lockdown, 7)provisions and dependence on others, 8)employment and income.

**Conclusions:** Parents and CYP were worried about the virus, although CYP less so. Parents and children however, were frustrated with the lack of specific and paediatric focused information and guidance, expressing disappointment with the adult centric information available. Parents also felt alone, especially with their concerns around the implications of cardiac service suspension and the implication for their child’s health. In order to better support children and their families, resources need to be developed to address families’ and CYP concerns for their health during this pandemic.

What is already known?

- People are feeling vulnerable and have heightened levels of stress during the COVID-19 pandemic
- Mental health and well-being of individuals has been adversely affected during this health crisis
- The COVID-19 pandemic has affected the mental health of parents due to uncertain financial circumstances, the closure of schools and suspension of educational services for children

What this study adds?

- Parents worry about the virus and are vigilant for symptoms of the virus or cardiac symptoms.
- Parents felt abandonded, unsupported by their Government, local and specialist CHD clinical team, and wanted information specfic to their child’s cardiac diagnosis
- Families feel forgotten and that medical/ surgical procedures children had been waiting for were no longer prioritized, which added another level of abandonment

## Background

In December 2019 a novel severe acute respiratory syndrome coronavirus 2 (SARS-CoV-2) was identified (1), and the subsequent rapid transmission of infection around the world resulted in the World Health Organization (WHO) declaring the outbreak of SARS-CoV-2 disease (COVID-19) a pandemic in March 2020(2). Despite the high infection rate amongst adults, children appear to be remarkably unaffected by COVID-19 (1-5% of cases)(3, 4), as well as experiencing milder disease and significantly lower mortality rate (4). In the first three months of 2020 governments of the majority of European countries put measures in slow the rate of infection, using social distancing and lockdown measures, lasting until early summer.

Congenital heart disease (CHD) represents one third of all major congenital anomalies, with a reported UK prevalence of 9 per 1,000 live births(5) resulting in approximately 5,000 CHD births per annum, of which 725 require interventional cardiac catheterisation and 4,000 need surgery per annum(6). The recent health crisis has seen an unprecedented cessation of many Health Service outpatient clinics and all elective surgical procedures, including cardiac surgery suspended for three months from March 2020 (7). The hiatus of usual paediatric cardiology services during this crisis period may have caused significant parental anxiety are well as unintended consequences due to delayed intervention(8). The aim of this study was to explore experiences, information and support needs, and decision-making of parents of a child with CHD in response to the COVID-19.

## Methods

A survey study of parents of children with CHD, children and young people (CYP); capturing experiences, decision-making, information and support needs during the COVID-19 crisis. The survey launched during the first infection wave of the COVID-19 pandemic within the UK and subsequent restriction of free movement under lockdown rules (opened 09/04/2020 and closed on 09/05/2020). The results in this study form part of a larger longitudinal study describing experiences of parents and children with various paediatric conditions. The study was approved by the University of Southampton and NHS Health Research Authority Research Ethics Committees (Ethics Number IRAS nr. 282176)

### Study participants

Parents of children and CYP with CHD were recruited through social media, national charities web pages and Facebook groups to minimise the burden on the health system during the COVID-19 outbreak. Electronic consent was obtained through the online survey. The aim was to recruit sufficient numbers of participants to describe the variety of concerns and experiences within closed statements(9) as well as common themes from the descriptive analyses of the rich qualitative data.

### Survey

The survey content is based on currently available literature(9, 10), expert health care professional input, and parents of children with CHD. Within the survey there a number of sections and number of closed statement items: Experiences(n=9), Information(n=4), Decisions(n=6) and Support needs(n=8). The closed statement items were on a Likert scale of Not at all(1), A little(2), Quite a bit(3), Very much(4). At the beginning of each section there was a free text box. The number of items was deliberately small, allowing for rapid analysis and dissemination. Feedback from parents (n=3) was sought about the survey in terms of content, language and inclusiveness of content, as well as the sensitivity around the timing of the research. Iterative changes were made accordingly.

### Data analysis

Descriptive statistics were completed using IBM Statistical Package for Social Science (SPSS) (version 25, IBM, Armonk, NY, USA) summarising demographic data, and simple descriptive statistics of the closed statement. A thematic content analysis was used to explore responses to the open text boxes, informed by a three-stage coding process (11, 12): stage 1). An initial sample of 34 comments were open coded into broad comment categories by two researchers (SS and RW), an initial framework was compiled, and conflict around themes was resolved with a third researcher (ASD); stage 2) the best-fit framework guided categorization of all comments from the data (LVM), with further refinement (SS and RW); stage 3) overarching themes were identified. The number of comments was counted to identify the weight of themes. As there was considerable overlap in the rich data within the sections, the total number of comments did not match the number of participants.

## Results

### Participants

#### Parents

184 people completed the survey, presented by 92%(n=169) mother, 9%(n=15) paternal/other. The median age of children was 8-years old (Interquartile range (IQR): 3, 13) and parents was 40(IQR: 33, 46). Children with CHD had a varying diagnosis with 16%(n=29) waiting for surgery and 84%(n=155) not waiting for any planned surgery. The geographic representation of respondents was well spread through the country with parents reporting from each of the four home nations of the UK (Table 1).

**Table 1:** Diagnosis.

| **Respondents – region** | **% (n=184)** |
| --- | --- |
| United Kingdom | 3.8% (7) |
| Northern Ireland | 20.1% (37) |
| Scotland | 5.9% (11) |
| Wales | 7.0% (13) |
| United Kingdom | 7.0% (13) |
| Southern England | 36.4% (67) |
| Northern England | 19.5% (36) |
| **Parent reported diagnosis** | **% (n=184)** |
| ASD, VSD or AVSD | 13.0% (24) |
| Pulmonary stenosis | 5.4% (10) |
| TOF or DORV | 17.4% (32) |
| Left heart obstruction | 14.7% (27) |
| Transposition of the great arteries | 11.4% (21) |
| Univentricular heart physiology | 26.1% (48) |
| Ebstein's anomaly | 0.5% (1) |
| Other complex CHD | 8.2% (15) |
| Cardiomyopathy or cardiac transplant | 2.2% (4) |
| Complete heart block | 1.6% (3) |
| Long QT syndrome | 1.1% (2) |
| **CYP reported diagnosis** | **% (n=36)** |
| Waiting for surgery | 2.7% (n=1) |
| No surgery planned | 80.5% (n=29) |
| Don't know | 16.7% (n=6) |
| VSD | 11.1% (4) |
| Pulmonary stenosis | 2.7 % (1) |
| TOF | 2.7% (1) |
| Left heart obstruction | 11.1 % (4) |
| Transposition of the great arteries | 16.6% (6) |
| Univentricular heart physiology | 13.8% (5) |
| Ebstein's anomaly | 2.7% (1) |
| Congenital heart disease/ don't know | 50% (13) |

ASD: atrial septal defect; AVSD: atrioventricular septal defect, CHD: congenital heart disease; CYP: children and young people; DORV: double outlet right ventricle; TOF: tetralogy of Fallot; VSD: ventricular septal defect

#### Children and young people

36 children and young people (CYP) completed the survey. The median age of CYP completing the survey was 18 years old (IQR:17,22). Thirty children indicated their age 43% (n=13) were <=18 years old and 57% (n=17) were >=18 years of age. CYP with CHD had a varying diagnosis with 2.7%(n=1) waiting for surgery and 72.2%(n=26) not waiting for any planned surgery (Table 1).

### Closed item statements

#### Parent

Many parents worry about the virus (86.4%), and potential symptoms (89%), in addition to considerable concern regarding the ability of their child’s heart to cope if cardiac symptoms were triggered by the virus (88%). Respondents felt they did not receive adequate/sufficient information from their child’s clinical team (85%), and wanted information specfic to their child’s cardiac diagnosis (82%). Over half of parents felt their child should be isolated from everyone except parents (54%) and worried about health care professionals (HCPs) coming into the home (70%). Parents were worried they might catch the virus (63%) and if so, their child would catch it from them (85%). All parents accessed information on social media (100%), which for some (48%) led to anxiety. Three-quarters of respondents worried hospitals were no longer a safe place (77%).Planned appointments or surgery (84%) were rescheduled in consultation with the medical team, with a proportion of parents (16%) making the decision not to attend without consulting their team (Figure 1). Parents would welcome support to reduce their own worries relating to the virus (36%) and help to support their children (28%).

**Figure 1:**
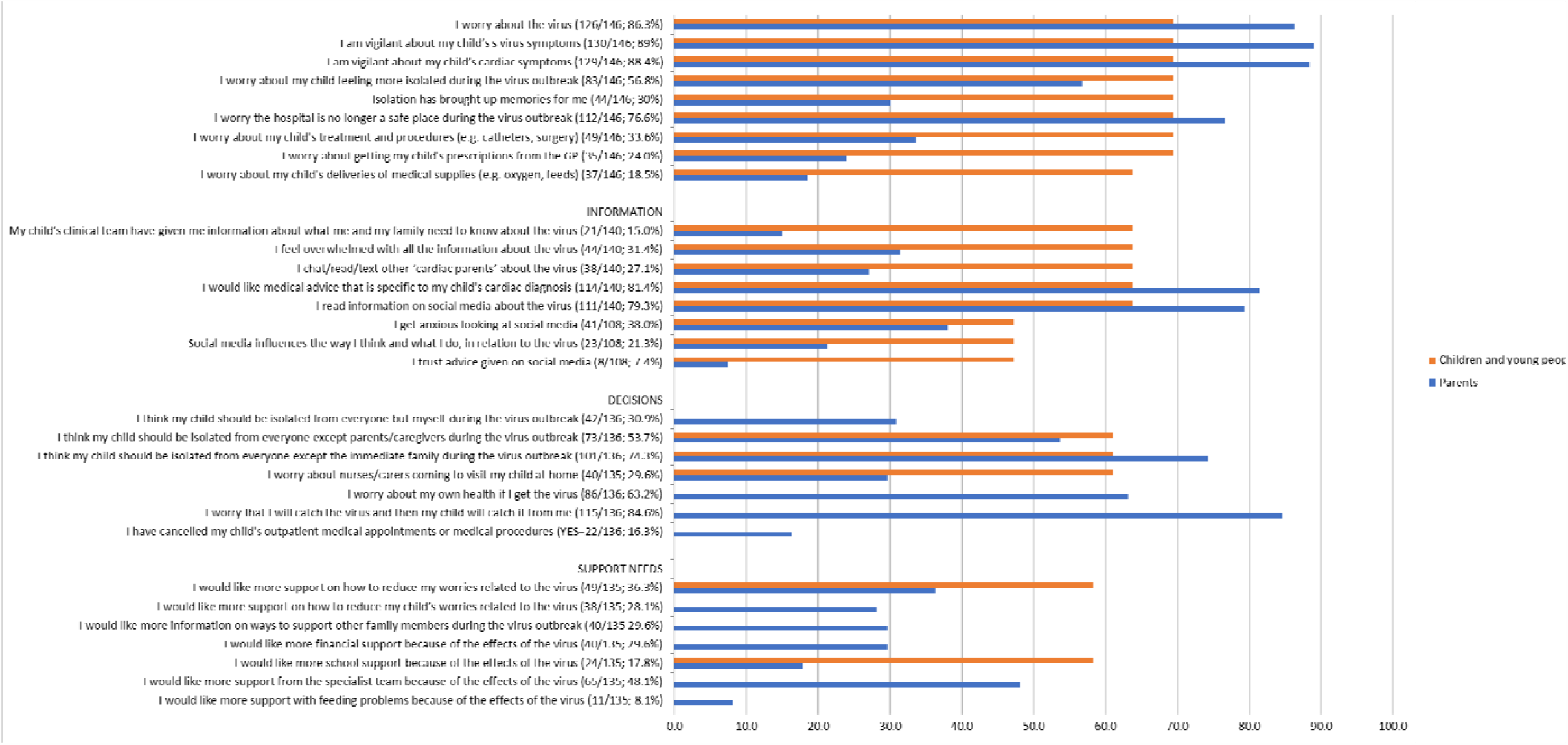
Children and young people and parent responses from closed statement item relating to; Experiences(n=9), Information(n=4), Decisions(n=6) and Support needs(n=8) represented as number and percentage of respondents

#### Children and young people

Worries of CYP mirrored that of parents, although to a lesser extent. CYP worry about the virus (69.4%), and are vigilant for symptoms of the virus (69%). The majority of respondents worried that hospitals were no longer a safe place (69%). Respondents felt they did not receive information they needed from their clinical team (64%), and would have wanted information specfic to their cardiac diagnosis (64%).

### Open text boxes

From the 184 parent respondents (n=141) provided comments, and from the 36 CYP respondents (n=25) comments. Overall, the responses covered 34 subthemes, forming eight overarching themes, related to the virus (four themes) and lockdown/shielding or isolation (four themes): *Virus:* 1)information, guidance and advice 2)change in health care provision, 2)risk of infection, and 4)fears and anxieties; *Lockdown and isolation:* 5)psychological and social impact, 6)keeping safe during lockdown, 7)provisions and dependence on others, 8)employment and income (Figure 2;Table 2).

**Table 2.**
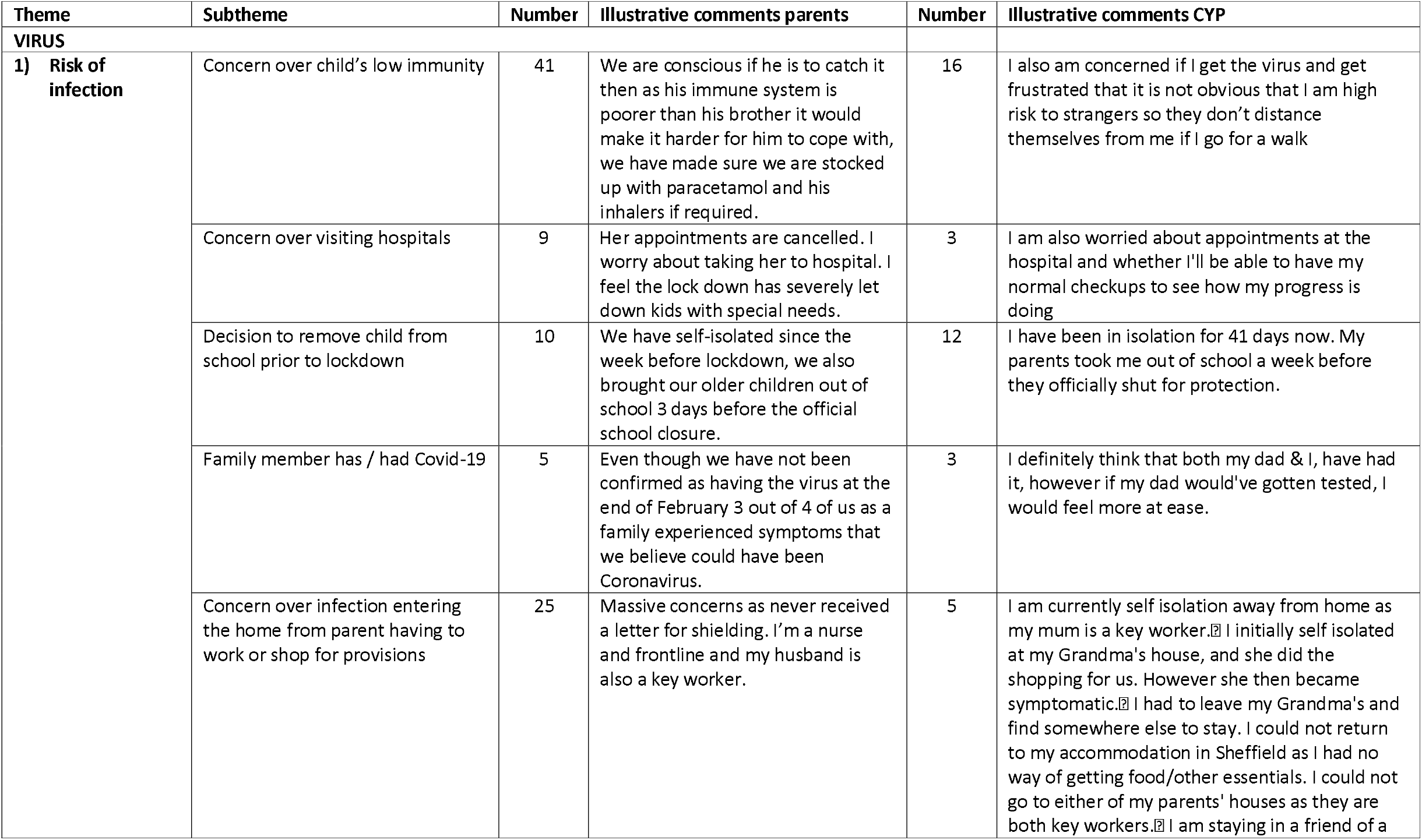
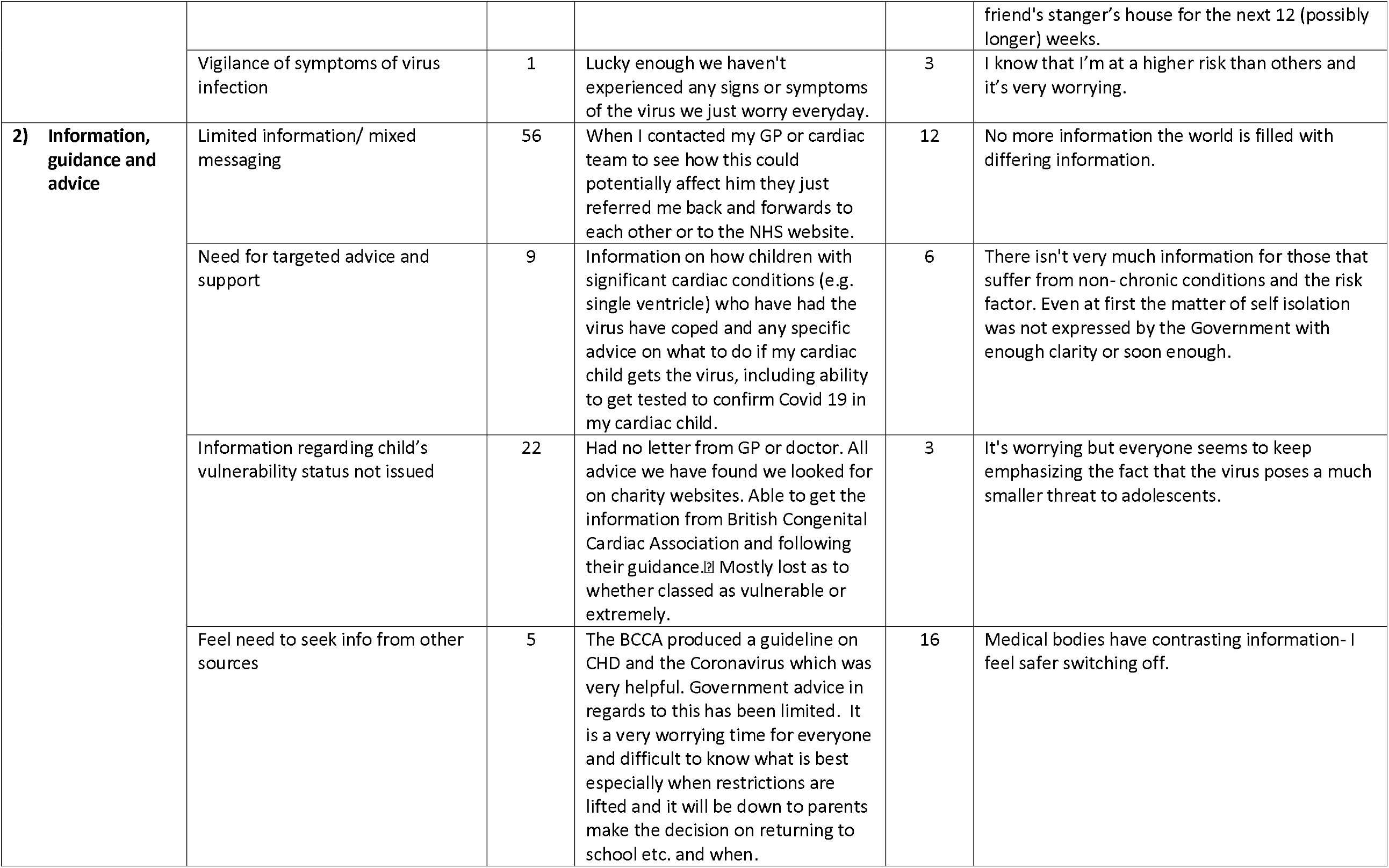
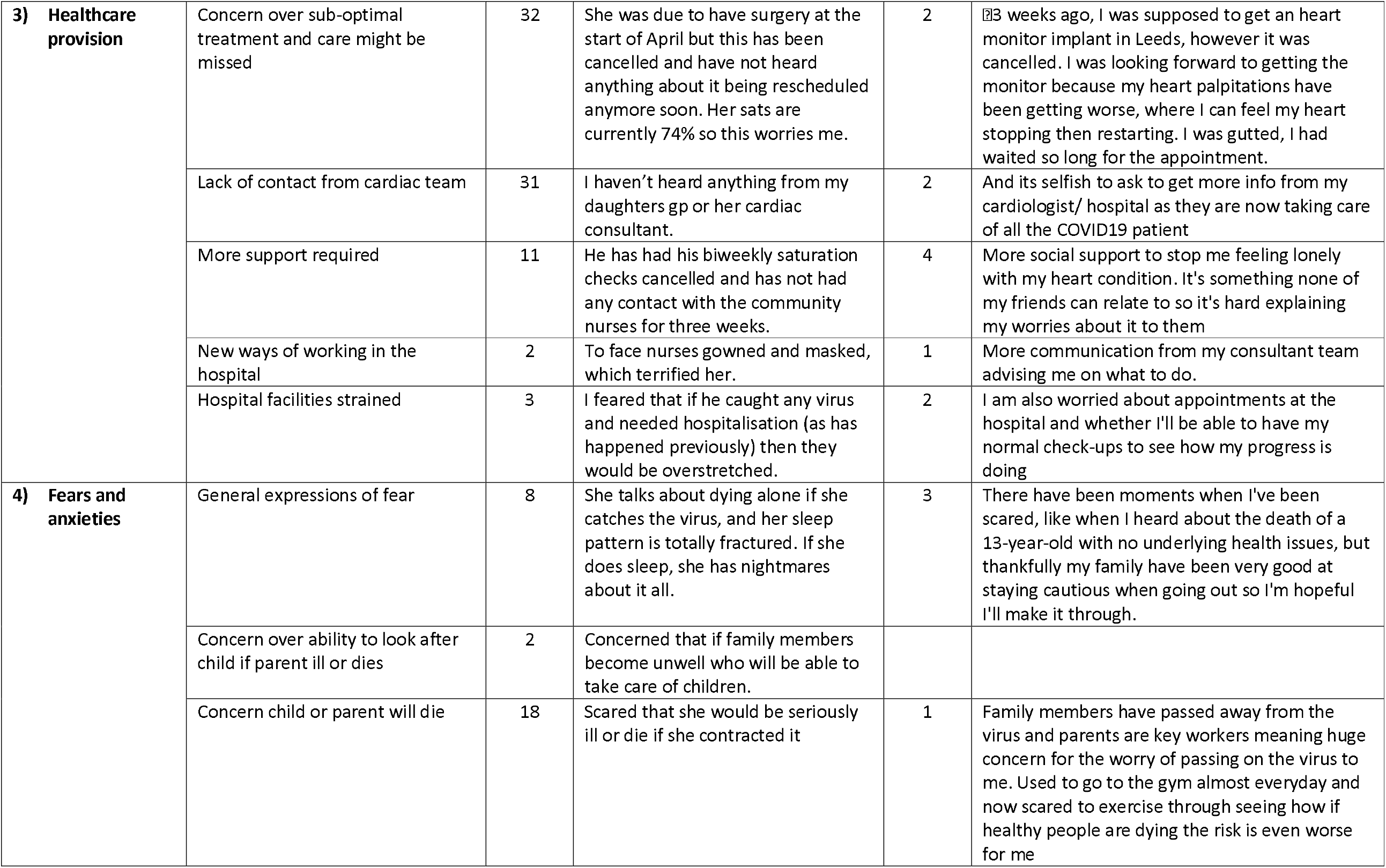
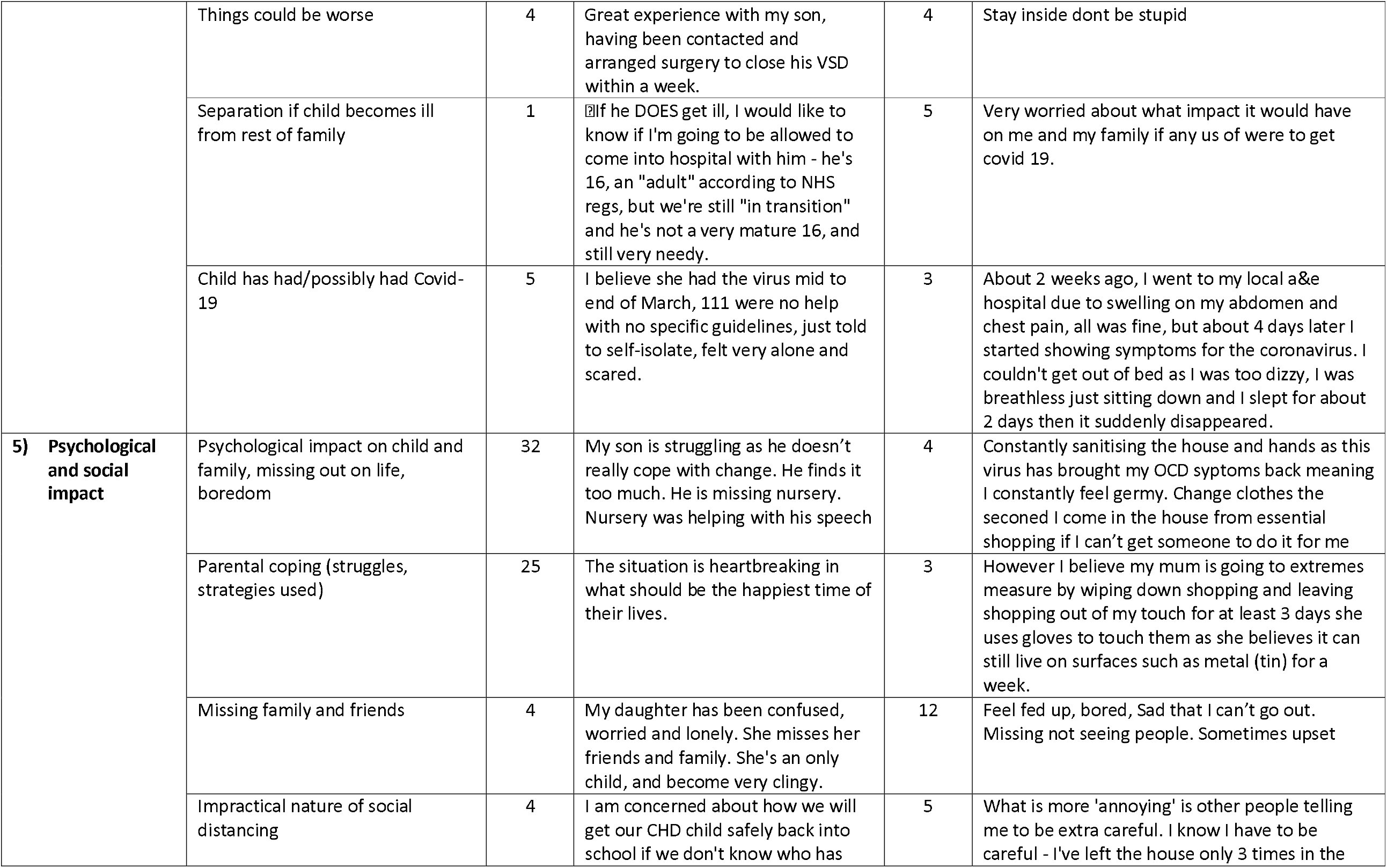
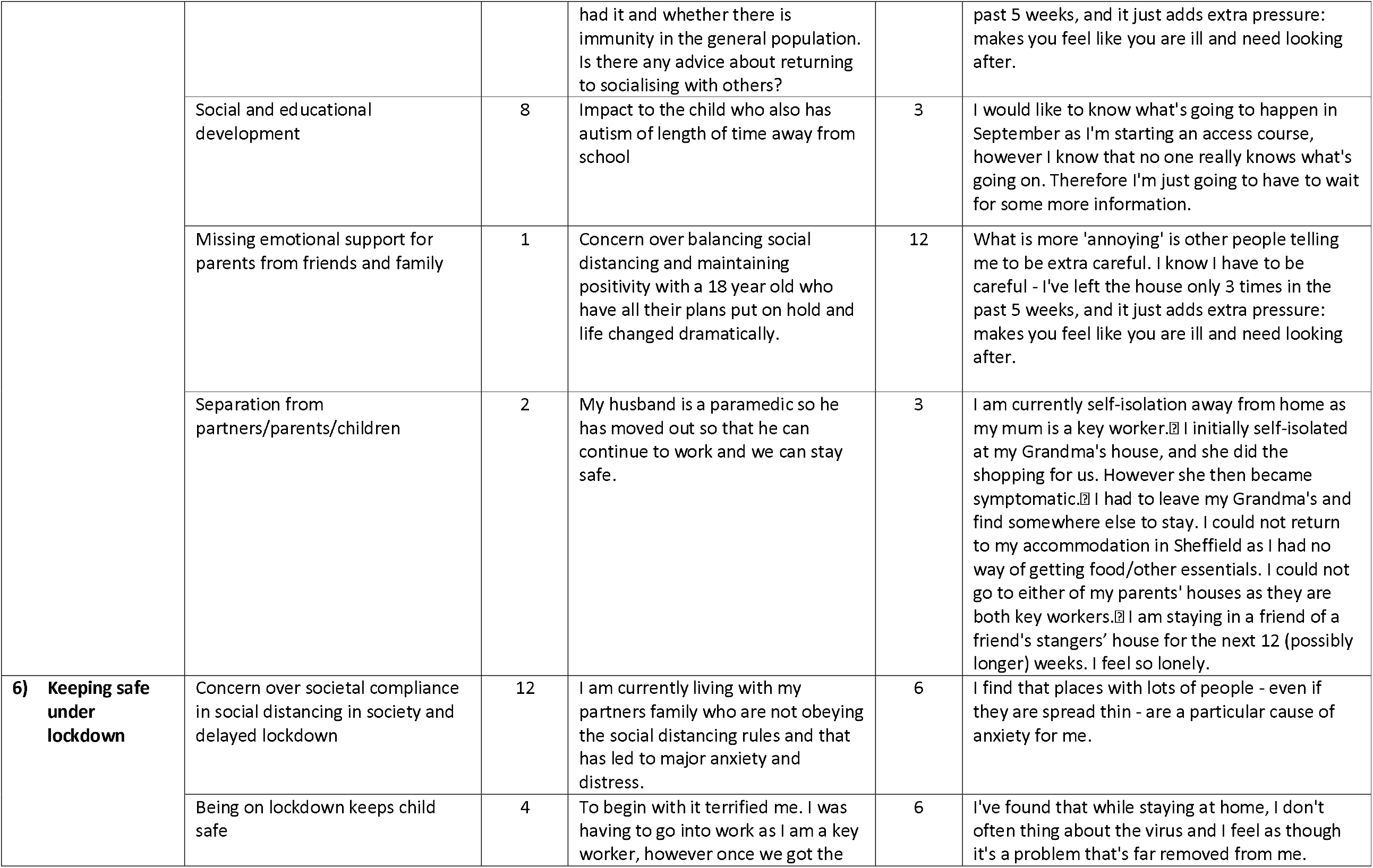
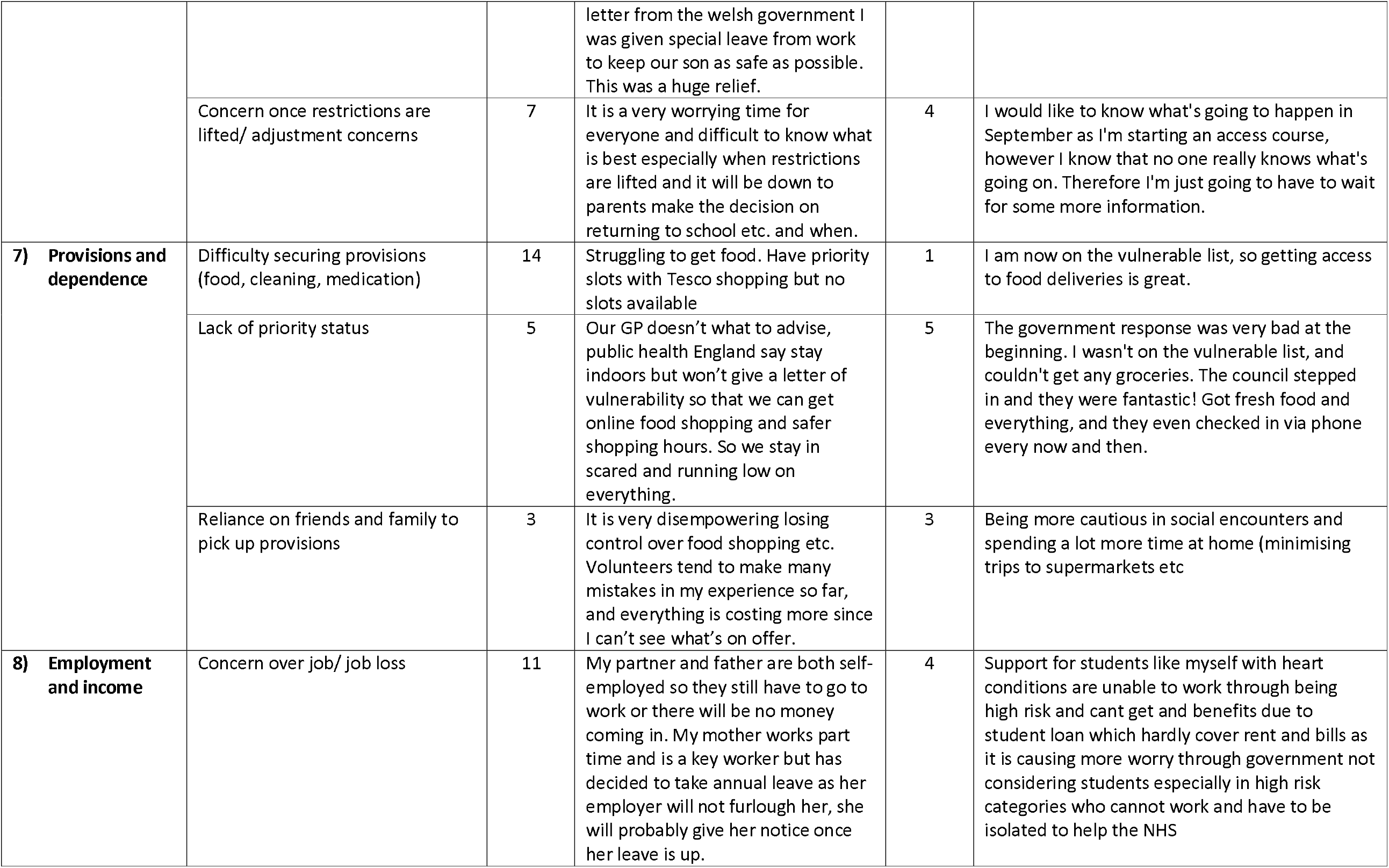
CHD: Experiences of Covid-19: Table of themes from 141 comments from a total of 184 parent respondents (76.6% of total respondents) and 24 comments from 36 CYP (66.6%)

**Figure 2:**
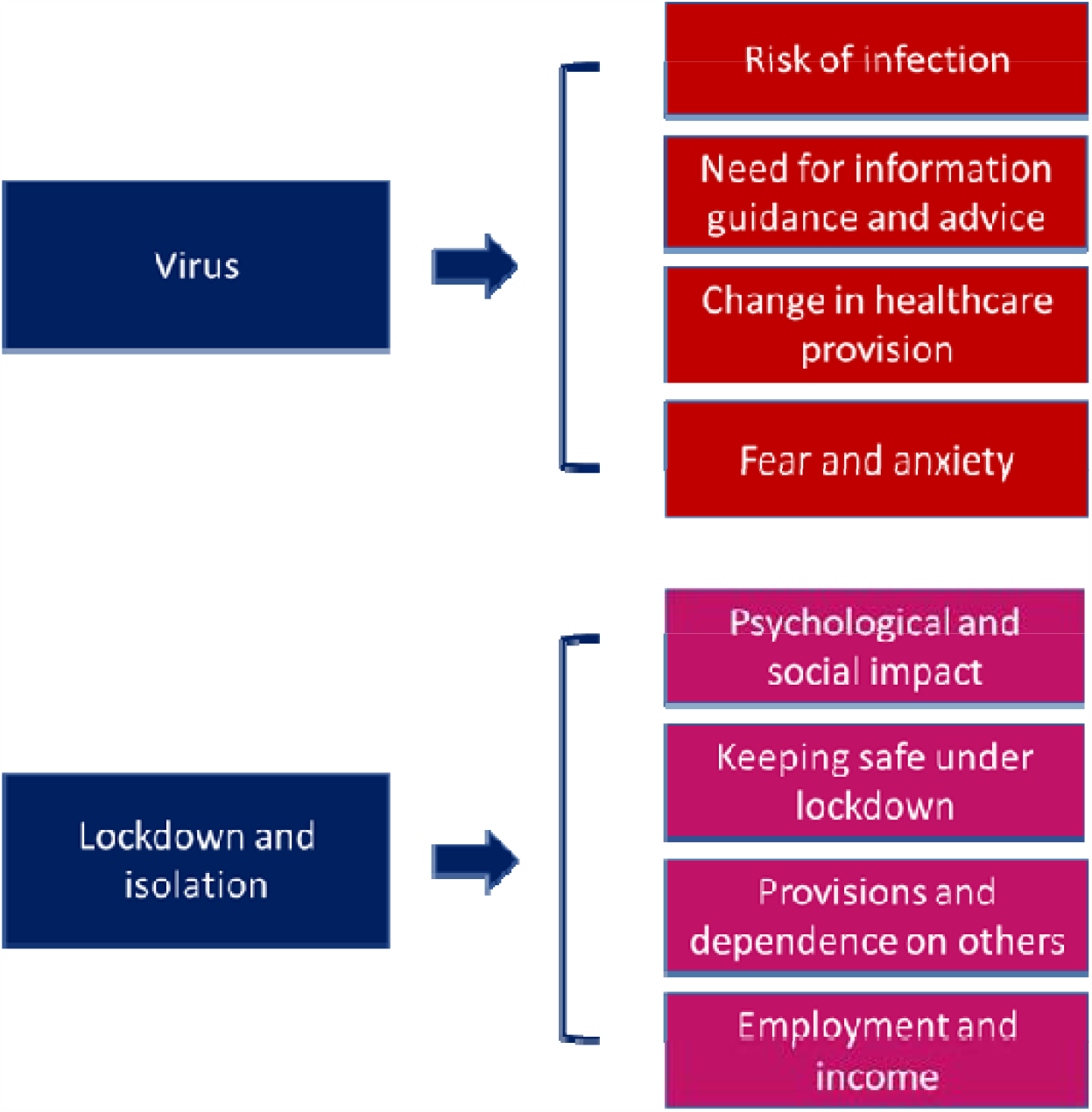
A comment model of parental experience of COVID-19 crisis.

### Virus

#### Parents

Most comments concerned the virus (n=56), with almost two-thirds of respondents (n=83) describing concerns about their child’s risk to their heart and ability to cope or overcome infection with the virus, how their heart or lungs would cope (n=18). Some parents (n=10) were worried about elective surgical procedures being delayed, and the negative consequences for their child. Many parents reported feeling abandoned by the specialist clinical team or the Government or HCPs in general (n=34).

Parents reported that information provided them was either non-existent or inconsistent within the home nations (n=21) they were also upset about the lack of support regarding the virus from their local team, specialist clinical team or The Government. Parents were frustrated the information provided related to adults. Parents wanted specific information relating to COVID-19 and their child’s particular heart defect, as well as what to do if they or their child contracted COVID-19. They as wanted reassurance they were not alone, and wanted letters to be able to stay home with their children (n=66).

#### Children and young people

Most comments from CYP described a fear of contracting the virus and not recovering (n=16), although some had no concerns (n=4). CYP wanted more information specific to their cardiac diagnosis (n=6) and almost half of them (n=12) had not left home since lockdown began. CYP would have liked more support with information specific to their cardiac diagnosis(n=7).

### Lockdown and isolation

#### Parents

Parents described the psychological impact of the emergence of the virus and subsequent lockdown (n=13), for both themselves and their children. Parents also raised concerns about keeping their child safe during lockdown, and how restrictions would be lifted to ensure safe re-integration. Accessibility of grocery deliveries and food supplies were a concern for parents(n=11). A small number of parents raised financial or employment worries(n=3), with some parents who worked on the frontline or were keyworkers lived separately to keep their family safe.

#### Children and young people

CYP reported feeling sad, panicked, isolated and missing their friends (n=12). The same number of children also reported feeling sad and bored.

### Information

#### Parents

Parents reported accessing most information from British Heart Foundation (n=15), other CHD charities (n=23) British Congenital Cardiac Association (n=9), in addition to news outlets (n=54), NHS/ Government website (n=44), hospitals (n=19), World Health Organisation (n=10) and social media (n=24).

#### Children and young people

Information sources were parents (n=8), news sites (n=16), although children reported worry and concern about conflicting/ untrustworthy information (n=6).

### Support

#### Parents

Parents wanted more support getting letters to gain delivery slots and what to do when lockdown restrictions are lifted (n=18).

#### Children and young people

CYP would have liked more support with mental health and well-being (n=4) and support with school or decisions around college.

### Decisions

#### Parents

The majority of parents (n=92) did not rely on official advice regarding clinical vulnerability and took the decision to keep their child at home ‘shielded’ based on ‘gut instincts’ and relying on their own judgment.

#### Children and young people

The majority of CYP described trying to keep safe by staying indoors, sanitizing hands and the family home (n=16). A number of children (n=4) were grateful for the study being conducted, as they felt they were often forgotten or overlooked.

## Discussion

To our knowledge this is the first study to report the experiences of parents of children and experiences of CYP with CHD during the COVID-19 crisis. The majority of parents expressed an overwhelming sense of worry about the effects of the virus, and the potential impact this may have on their child’s heart should they contract the illness. This worry also resonated amongst CYP, although fewer were as concerned about the virus and some children were able to rationalize that adolescents did not appear to be much affected by the virus.

The majority of parents surveyed electively withdrew themselves and their children with CHD and other family members from society before or immediately as government lockdown was initiated. Reasons for this were fear of the unknown effects arising from a lack of specific information, and distrust in the adult-centric advice given. This was similarly echoed by CYP who wanted paediatric-specific information. Some of the children and CYP included in this survey had unrepaired or palliated CHD, but many had haemodynamically inconsequential lesions, and it is striking that even these families experienced uncertainty and anxiety related to their CHD-associated risks.

Although clinical teams provided support to parents and CYP with CHD, providing available information during the COVID-19 crisis, this is not how it was perceived by parents, particularly with respect to letters to support them being able to stay at home with their children. Further to this, parents felt abandoned and alone, especially with their concerns around the implications of cardiac service suspension and the implication for their child’s health. Concerns regarding perceived vulnerability of their children may have been further exacerbated following reports of COVID-19 or COVID-19 like symptoms with hyperinflammatory multisystem syndrome temporally associated with COVID-19 (PIMS-TS) and atypical Kawasaki disease(13), as such it may be an important lesson for us as a health care community to recognize the potential psychological impact media reports may have on parents of children with CHD(14), and their increased vulnerability as a result.

CYP reported feeling panicked, isolated and missing their friends, as well as feeling sad and bored. There are profound consequences for the mental-health and well-being of children as a result of social distance strategies to contain the spread of the virus(15); in addition there may be long-lasting consequence of educational poverty and ‘diminution of educational opportunities’ arising from school-closures(16, 17), which may more adversely affect children with serious conditions. This pandemic has raised many societal issues relating to how children are treated and viewed during a pandemic(17), and the interests of children, particularly those with complex health and social needs, should be at the heart of any recovery plan to get back to normal or ‘new normal’. Larcher *et al*(17), raised an important question, ‘*how society views children; should they be regarded as pawns, pathfinders or partners in this enterprise’*. We assert that this process starts with providing children with specific information relating to the effects of COVID-19 on children as a whole, as well as in the context of those with serious conditions. Our survey suggests parents of children with CHD, as well as CYP feel scared, abandoned and frustrated at the lack of bespoke information that talks to them about them during the COVID-19 crisis.

The limitations of this work include the small sample size and potential exclusion of individuals with digital poverty, literacy and language issues. Although charities and support groups of children with CHD were targeted, some groups might have been overlooked thus imposing sample bias. The survey also represents a maternal view-point as the majority of respondents in the adult survey were women.

Despite these limitations we believe the findings of this survey are important requiring a national conversation as to how parents and their children with CHD are better supported in the future, particularly if further periods of social distancing measures are required which result in closure of schools. Larcher et al(17) propose that article 12 of the United Nations Convention on the Rights of the Child(18) should be invoked requiring, ‘*children to be informed and consulted over matters that concern them and that their views be given due weight in accordance with their age and maturity’* and is in keeping with the NHS ethos of ‘*no decision about me, without me’*. For the future, it is imperative we ensure better support and information for families to reduce the effects of social isolation, medical and educational deprivation.

## Conclusion

Parents were around the effects of the virus, as were CYP although to a lesser extent. Parents felt unsupported, abandoned and frustrated with the lack of specific and focused information and guidance, expressing disappointment with the adult centric information. In order to better support children and their families, developed resources need to address families’ concerns for their children’s health during this pandemic.

## Data Availability

None

## ACKNOWLEDGEMENTS

To parents and children with congenital heart disease for taking the time to share their experiences of the health care crisis as a result of COVID-19, to the British Heart Foundation, Children’s Heart Federation and regional CHD charities for sharing the information via social media and websites.

## CONTRIBUTORS STATEMENT

Authors made the following contribution to the manuscript: (1) Anne-Sophie Darling formulated the original idea and wrote the study design including the survey, (2) Luise Marino, Richard Wagland, Sam Sodergren, Anne-Sophie Darlington, David Culliard analysed the data, (3) Luise Marino drafted the manuscript, (4) Tara Bharucha, Richard Wagland, Sam Sodergren, Anne-Sophie Darlington contributed to revising the manuscript for important intellectual content, (4) and all authors provided final approval of the version to be submitted.

